# Visuospatial processing impairment following mild COVID-19

**DOI:** 10.1101/2021.02.18.21251442

**Authors:** Jonas Jardim de Paula, Rachel Elisa Rodrigues Pereira Paiva, Danielle de Souza Costa, Nathália Gualberto Souza e Silva, Daniela Valadão Rosa, José Nélio Januário, Luciana Costa Silva, Débora Marques de Miranda, Marco Aurélio Romano-Silva

## Abstract

Severe Acute Respiratory Syndrome Coronavirus 2 infection causes coronavirus disease 2019. COVID-19 was an unknown infection that reached pandemic proportions in 2020 and has shown to bring long-term negative consequences. Here, we used a case-control design to investigate the performance of relatively young people recovered from COVID 19 in objective neuropsychological tests. We found significant differences between groups for all measures of the ROCFT with a large difference in the copy, a moderate difference in immediate recall, and a large difference in delayed recall. No significant differences were found for the measures from all the other five neuropsychological tests used.About one quarter of COVID 19 patients were below the 10th percentile according to normative data.

## Introduction

Severe Acute Respiratory Syndrome Coronavirus 2 (SARS-CoV-2) infection causes coronavirus disease 2019 (COVID-19). COVID-19 was an unknown infection that reached pandemic proportions in 2020 and has shown to bring long-term negative consequences (Wang, Kream & Stefano, 2020). People recovered from COVID-19 may still present complications including respiratory and neurological manifestations (Lopez-Leon et al., 2021). In other neurotropic viral infections, cognitive sequelae may occur due to brain damage or dysfunction owing to vascular lesions and inflammatory processes (Levine, Sacktor & Becker, 2020). Persistent cognitive impairments from viral infections are associated with activities of daily living and psychosocial adaptation (de Paula et al., 2020). Much from COVID-19 consequences is still veiled from its novelty. Here, we used a case-control design to investigate the performance of relatively young people recovered from COVID-19 in objective neuropsychological tests.

## Method

We assessed 49 patients who had recovered from mild COVID-19, meaning they had flu-like symptoms, but no need for hospitalization, mechanical ventilation or any oxygen therapy support. Infection was confirmed by polymerase chain-reaction assay. All participants gave written consent. The study was approved by the local ethics board (registry: 3768820.1.0000.5149).

A control group of 49 participants was selected from a sample assessed with neuropsychological testing from 2017 to 2019. We adopted a quasi-experimental design matching each patient by sex and formal education. Post COVID-19 and control groups were composed of 38 female and 11 male participants each. Both groups had two participants with less than 9 years of education, 13 with a high-school degree, and 34 with at least a college degree. No significant difference (*p*=0.505) was found for age between post-COVID-19 (35.9±9.7 years) and control (34.6±10.8) groups.

Neuropsychological assessment of the post-COVID-19 group was carried out before participants’ neuroimaging and took about 40 minutes. A certified neuropsychologist and clinical or research assistants under his direct supervision conducted the assessments. Cognitive tests done were the Rey-Osterrieth Complex Figure Test (ROCFT) (copy, immediate and delayed recall), Category Fluency (animals, fruits, and a switching version), Trail Making Test (Parts A and B), Five Points Test (180 seconds), Digit Span (forward and backward), and the Logical Memory Test (Immediate and Delayed Recall).

**Figure 1.**
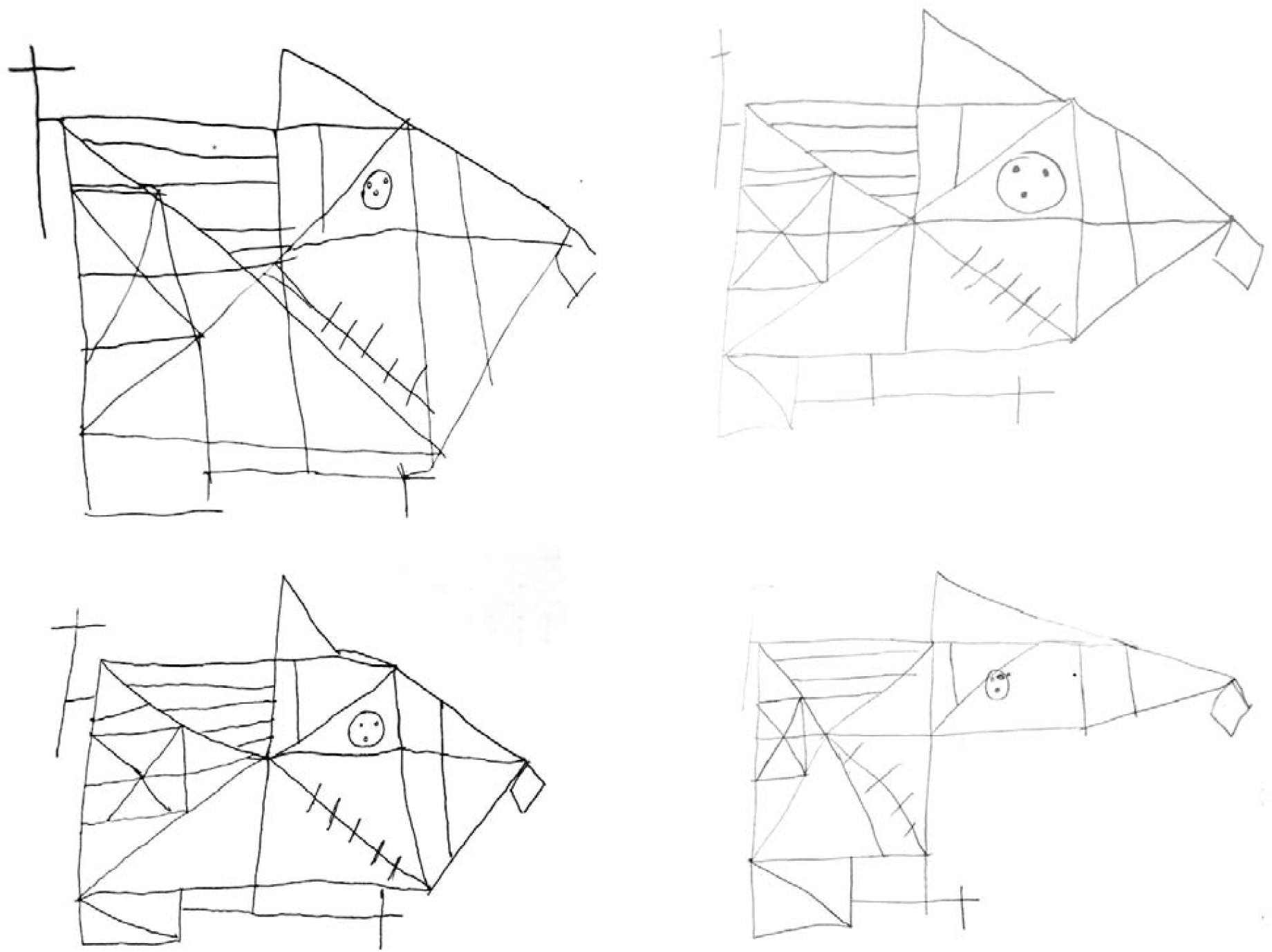
Performance of four recovered mild COVID-19 patients in Rey-Osterrieth Complex Figure test (copy)

## Results

Results are shown in Table 1. We found significant differences between groups for all measures of the ROCFT with a large difference in the copy (d=1.31), a moderate difference in immediate recall (d=0.58), and a large difference in delayed recall (d=0.82). No significant differences (p>0.05) were found for the measures from all the other five neuropsychological tests used.

**Table 1:**
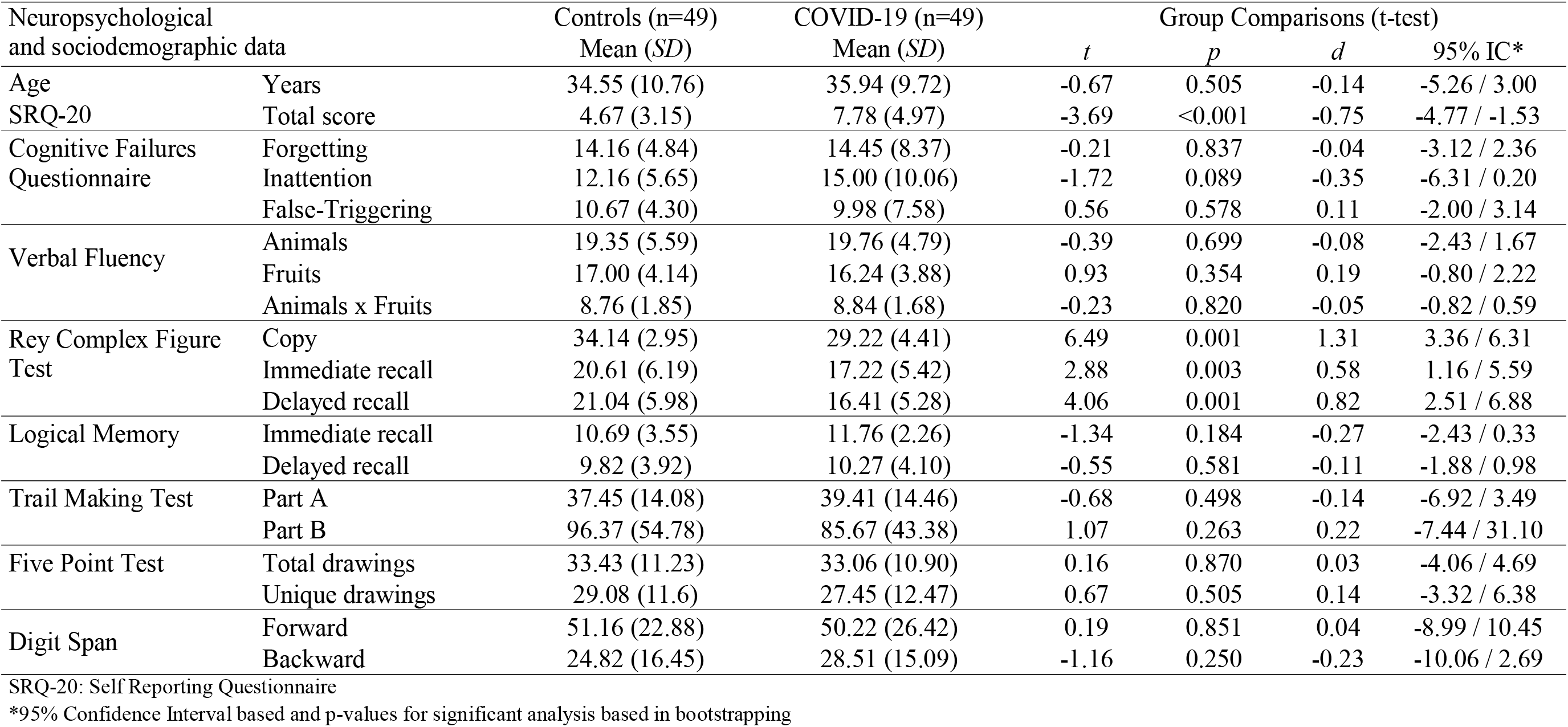
Participants description and comparison of mild-COVID-19 recovered patients and controls on neuropsychological and sociodemographic measures.

Classifying ROCFT copy scores according to Brazilian normative data 12 (24%) COVID-19 patients were classified as inferior (below the age-corrected 10th percentile), against 2 (4%) of the control group. For the immediate memory only 1 patient (2%) and 2 controls (4%) performed in this range.

## Discussion

We provided preliminary evidence of visuospatial impairment in patients who recovered from mild COVID-19. Immediate and delayed deficits in the ROCFT copy and recall could not be explained by socio-demographic factors or emotional symptoms, suggesting a neurocognitive deficit caused by SARS-COV-2 infection. The performance on visuoconstruction and memory tests, such as the ROCFT, are associated with different aspects of everyday life, including the capacity to learn, problem-solving skills, and activities of daily living (Davies et a., 2011)

Considering the relatively high prevalence of cognitive deficits in neuroinfections (Levine et al., 2020) and their impact on daily living, it is paramount to investigate impairments caused by COVID-19. Although we must consider pulmonary or neurovascular damage, other mental disorders, and psychosocial factors (Wang et al., 2020; Lopez-Leon et al., 2021; de Paula et a., 2020) as contributing factors, future studies should address the etiology of cognitive symptoms as a direct consequence of SARS-CoV-2 neuroinfection.

## Data Availability

Data is restricted by the local ethics Board

## Acknowledgments

Grants: Coordenação de Aperfeiçoamento de Pessoal de Nível Superior -CAPES -Brazil; Conselho Nacional de Desenvolvimento Científico e Tecnológico -CNPq -Brazil. We would like to thank Dr. Danielle de Souza Costa for helping with RCFT correction and interpretation.

